# WORKING MEMORY FILTERING AT ENCODING AND MAINTENANCE IN HEALTHY AGEING, ALZHEIMER’S AND PARKINSON’S DISEASE

**DOI:** 10.1101/2024.06.18.24309107

**Authors:** Sofia Toniolo, Robert Udale, Verena Svenja Klar, Maria Raquel Maio, Bahaaeddin Attaallah, George Tofaris, Michele Tao-Ming Hu, Sanjay George Manohar, Masud Husain

## Abstract

The differential impact on working memory (WM) performance of distractors presented either at encoding or during maintenance was investigated in Alzheimer’s (AD), Parkinson’s Disease (PD) and healthy ageing. Across three studies, 28 AD and 28 PD patients, 28 elderly (EHC) and 28 young healthy controls (YHC) were enrolled. All participants performed a delayed reproduction task, where they reported the orientation of an arrow from a study set of either two or three items, with a distractor present either at encoding or at maintenance. Mean absolute error (the difference between probed and reported orientation) was calculated as an analogue measure of WM. Additionally, mixture model metrics i.e., memory precision, target detection, misbinding (swapping the features of an object with another probed item) and guessing were computed. MRI data was also acquired in AD, PD and EHC participants, and whole hippocampal volumes were extracted to test whether WM filtering and overall performance were related to hippocampal integrity.

EHC and PD patients showed good filtering abilities both at encoding and during maintenance. However, AD patients exhibited significant filtering deficits specifically when the distractor appeared *during maintenance.* Healthy ageing and AD were associated with higher rates of both misbinding and guessing, as well as lower target detection, and memory precision. However, in healthy ageing there was a prominent decline in WM memory precision, whilst in AD lower target detection and higher guessing were the main sources of error. Conversely, PD was associated only with higher guessing rates. Hippocampal volume was significantly correlated with filtering during maintenance – but not at encoding – as well as with overall mean absolute error, target detection, guessing and misbinding. These findings demonstrate how healthy ageing and neurodegenerative diseases exhibit distinct patterns of WM impairment, including differential effects on filtering irrelevant material presented at encoding and maintenance.

## 1. Introduction

Working memory (WM) is a limited data processing system responsible for encoding and briefly maintaining information, so that it can still be accessed in the absence of inputs from the environment.^1,2,3^ Encoding processes transform transient perceptual representations into a more durable state, while maintenance mechanisms allow information to be retained so that it can be used to guide behaviour over brief delays.^4,5^ WM performance may decline with ageing,^6,7,8^ and is further impaired in several neurological conditions such as Alzheimer’s Disease (AD)^9,10^ and Parkinson’s Disease (PD).^11,12^ In *healthy ageing*, several components contributing to declining performance in visual WM have been identified: a reduction in the number of items that can be stored, or WM capacity^13^; a deficit in maintaining the associations (bindings) between individual object features^14^ resulting in a higher proportion of misbinding or “swap errors”^15^; an increase in random guessing^16,17^; and a decrease in response precision.^18,19^ Since WM is limited in capacity, items compete for access to it. Hence, the ability to efficiently *filter* out irrelevant information is crucial for optimal use of WM resources, and its impairment constitutes an additional source of decline of WM performance and capacity.^20,21,22,23,24,25,26,27^

Filtering out distractors at *encoding* (i.e., when the items are presented), versus *maintenance* (i.e., when they are retained in memory), poses different challenges.^28,29^ Filtering at encoding has been envisaged as deployment of a flexible gating system, allowing at the same time relevant targets to enter WM while keeping distractors out.^29,30^ Filtering during maintenance on the other hand might rely on the ability to keep the gate shut firmly once targets are successfully encoded, in order to prevent any further information entering WM and corrupting task-relevant information.^24,26,27,30^ The integrity of fronto-striatal networks has been commonly associated with performance in filtering paradigms,^31,32,33^ and linked to brain structures involved in WM storage including the parietal cortex.^31,34^ However, the contribution of the hippocampus to filtering abilities, despite being crucial for short-term and long-term memory function,^35,36,37,38^ has received little attention.

Elderly participants seem to be particularly prone to errors with increasing memory load,^13^ which has been linked to a decline in their filtering abilities.^20^ Some evidence suggests older adults show poorer performance if distractors are presented during maintenance rather than at encoding.^29^ It is currently unknown whether this is due to decreased memory fidelity, to a failure in successfully form a bound, stable representation of an object, or to information exceeding WM capacity, leading to random guessing. These questions can be addressed using delayed reproduction paradigms that ask participants to reproduce from memory, using a continuous analogue response space, specific features of items presented at encoding.^2^ Both an age-related decline in memory precision^18,19^ as well as increased guessing^39^ have previously been reported using such paradigms. Evidence of increased misbinding has been more mixed, with some data showing higher rates in healthy ageing,^15^ while other have not.^40^

Maintenance of bound features seems to be even more impaired in AD compared to healthy ageing.^41^ Increased misbinding in patients with both sporadic, late-onset AD and familial AD has now been reported using both change-detection and delayed reproduction tasks,^12,35,42,43,44^ and specifically linked to hippocampal dysfunction.^45,46,47^

An increased vulnerability to distractors is also detectable from the earliest phases of AD,^48,49,50,51,52^ particularly if task instructions need to be maintained over time, pointing towards a deficit of goal-maintenance processes.^53,54^ Crucially, comparative data on distractor filtering presented at encoding or maintenance within the same study are lacking.

PD is also associated with deficits in visual WM. Different paradigms have provided evidence of reduced WM capacity^55^ as well as greater levels of guessing, rather than misbinding.^12,56^ Further, PD patients have been shown to exhibit deficits in distractor filtering at encoding, which might be linked to basal ganglia and prefrontal dysfunction.^31,55^ However, other studies have shown that PD patients also fail to efficiently manipulate information over the maintenance period, which can be selectively modulated by replenishing their dopaminergic tone.^17^

In summary, healthy ageing, AD and PD have been associated with reduction of filtering abilities, which impact negatively on their overall WM capacity and performance. Comparative data on filtering at encoding and maintenance in these groups are currently lacking. Here, using a unified delayed reproduction task and computational modelling, we investigated whether filtering deficits in healthy ageing, AD and PD are related to a decline in memory precision, a decreased ability to detect the correct target, an increased likelihood of misbinding or greater random guessing. We also examined whether these performance metrics relate to hippocampal integrity, given the crucial role of the hippocampus in both encoding and maintenance processes.^36,57,58^ First, we established normal performance in young and elderly healthy controls (Study 1). Then, we studied filtering ability in AD (Study 2) and PD (Study 3).

## 2. Study 1 | Effects of healthy ageing on working memory filtering

### 2.1 Materials and methods

#### 2.1.1 Participants

28 young (YHC) and 28 elderly healthy controls (EHC) were recruited (YHC: 18-35 years and EHC: 50-90 years), through respectively either the departmental online participant recruitment system (SONA), or open day events. All performed the Filtering at Encoding and Maintenance Task (**Figure 1**). We also collected a measure of global cognition – the Addenbrooke’s Cognitive Examination-III (ACE-III)^59^ – in the elderly group as baseline screening for cognitive impairment. Participants who reported any psychiatric or neurological illness, were on psychoactive drugs or scored below the cut-off for normality (88/100 total ACE-III score) were excluded from the study. A summary of participants’ demographics and tests results is presented in **Supplementary Table 1**.

**Figure 1.**
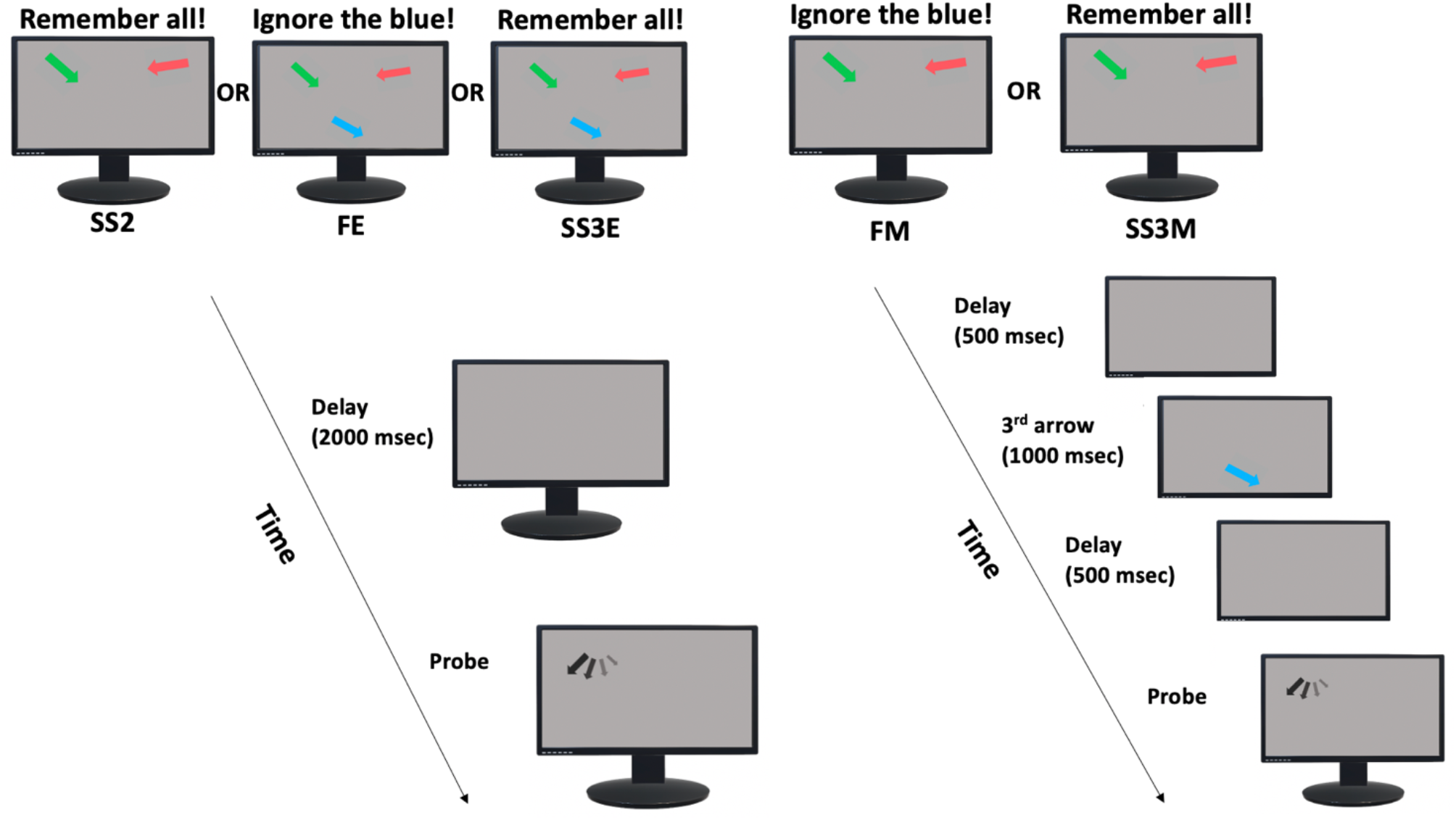
| Filtering at Encoding and Maintenance Task design. Participants were presented with either two (SS2) or three (SS3E) arrows at encoding and were instructed to remember the orientations of all arrows **(SS2 and SS3E)**, or to ignore one of them in the **Filter at Encoding (FE)** condition. After a blank interval (2000 msec), one of the target arrows (probe) re-appeared in black in its original location but in a random orientation, which the participants had to rotate to its remembered orientation. In the **Filter at Maintenance (FM**) and **Set Size three at Maintenance (SS3M)** the first two arrows appeared simultaneously and had to be remembered, and after a 500 msec delay during the maintenance period, a third arrow appeared for 1000 msec, which either had to be ignored (FM), or remembered (SS3M). After 500 msec the probe was displayed and had to be rotated to its original orientation.

For all three studies, the following principles were observed: All participants had normal or corrected-to-normal vision acuity and no color blindness. All participants gave written informed consent prior to the start of the study and received financial compensation for their participation. Ethical approval was granted by the University of Oxford Medical Sciences Inter-Divisional Research Ethics Committee (MS IDREC).

#### 2.1.2 Filtering at Encoding and Maintenance Task

The Filtering at Encoding and Maintenance Task (**Figure 1**) had 5 different conditions.

In **Set Size 2 (SS2)** and **Set Size three at Encoding (SS3E)** conditions, participants were presented with either two (SS2) or three (SS3E) different coloured arrows (red, green or blue) and were instructed to remember their colours and orientations. After a blank delay interval (2000 ms), one of the target arrows (i.e., the probe) re-appeared in black in its original location but presented in a random orientation. Participants were instructed to rotate the probe using the left and right arrow keys on a keyboard back to the remembered orientation. In the **Filter at Encoding (FE)** condition three arrows were presented simultaneously, as in the SS3E condition, but now participants were asked to ignore one of them.

In the **Filter at Maintenance (FM)** and **Set Size three at Maintenance (SS3M)** conditions, the first two arrows appeared simultaneously and had to be remembered. Subsequently, after a 500 msec delay during the maintenance period, a third arrow appeared for 1000 msec, which either had to be ignored (FM) or remembered (SS3M) and after 500 msec the probe was displayed. Further task specifics can be found in Supplementary materials.

#### 2.1.3 Statistical analysis

All analyses were conducted using MATLAB 2019a and JASP (JASP team, 2020). As first step, we computed the ***mean absolute error* (MAE)** for each condition for each participant. This provides an index of raw error. We then fitted the probabilistic Mixture Model introduced by Bays et al in 2009^60^ to each participant for each condition. The mixture model allows modelling of responses in terms of probability of correctly identifying the target (**target detection**), misbinding the features of an object with another among the probed items (**misbinding or “swap errors”**), random guessing (**guessing**), and extracts a measure of memory **precision**, (**Supplementary Model 1**).^60^

##### Key parameters used for statistical analyses

For all variables, **Instruction x Condition x Group** frequentist mixed ANOVAs were performed, where:

- **Instruction** – Refers to 3-item conditions with filtering (FE, FM) vs no filtering (SS3E, SS3M)
- **Condition** – All stimuli at encoding (FE, SS3E) vs one presented at maintenance (FM, SS3M)
- **Group -** YHC vs EHC

We also computed from the MAE a main effect of **Set Size**, both **at Encoding** and **at Maintenance** and their difference or **Set Size Cost**:

- **Set Size at Encoding =** SS3E - SS2
- **Set Size at Maintenance =** SS3M - SS2
- **Set Size Cost =** Set Size Encoding - Set Size Maintenance

A main effect of Filtering, or **Filtering rate**, was calculated separately at Encoding and Maintenance from MAE as follows:

- **Filtering rate at Encoding =** FE - SS2
- Filtering rate at Maintenance = FM - SS2
- **Filtering rate Cost =** Filtering rate at Encoding - Filtering rate at Maintenance

If perfect filtering was achieved, Filtering rate would be zero. The higher the Filtering rate, the worse the performance. For the ANOVA, effect size was quantified using Eta Squared (η2). Between-group differences for Set Size and Filtering rate metrics were assessed either with a t-test or Mann-Whitney U test, and a Bayesian independent sample analysis was performed. For the Bayesian analysis, we used the default priors for the effects (Fixed effects: r = 0.5, Random effects: r = 1).

### 2.2 Results

#### 2.2.1 Mean absolute error (MAE)

EHC performed significantly worse than their younger counterparts across all conditions (main effect of Group [F (1,48) = 40.46, p < 0.001, η2 = 0.292]; Figure 2A). Furthermore, both groups were significantly worse in the conditions where all three items had to be remembered (SS3E and SS3M) compared to when they had to filter an item out (main effect of Instruction [F (1,48) = 17.75, p < 0.001, η2 = 0.049]). There was no significant main effect of Condition, suggesting that overall Encoding conditions were not more cognitively taxing than Maintenance conditions. However, there was a significant 3-way Group x Instruction x Condition interaction [(F (1,48) = 8.22, p = 0.006, η2 = 0.014], suggesting EHC performed significantly worse if the three items to be remembered were presented simultaneously at encoding, compared to maintenance.

**Figure 2.**
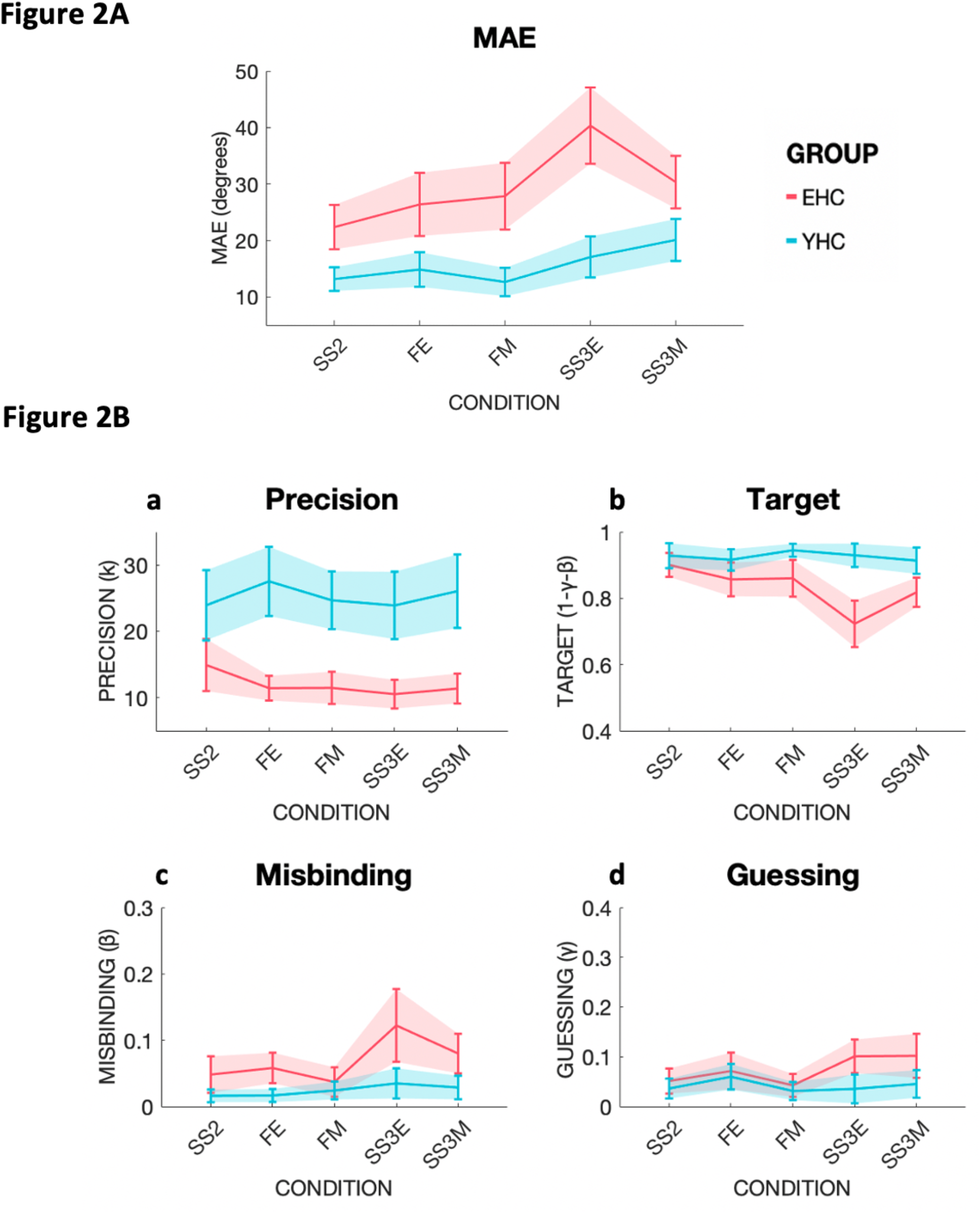
| Mean absolute error (MAE) and mixture model metrics in young (YHC) and elderly healthy controls (EHC) Performance of participant groups are shown labeled with different colors, EHC in coral red, YHC in turquoise. Set Size 2 (SS2), Filtering at Encoding (FE), Filtering at Maintenance (FM), Set Size three at Encoding (SS3E), Set Size three at Maintenance (SS3M). **2A:** MAE performance across the five different conditions. On the Y-axis MAE in degrees. Shaded areas represent confidence intervals (CI). **2B:** Mixture Model metrics across conditions in YHC and EHC. Panel a: Precision as concentration parameter κ, Panel b: Target probability, panel c: Misbinding probability, panel d: Guessing probability.

#### 2.2.2 Impact of Set Size at Encoding and Maintenance

EHC exhibited a higher Set Size effect at Encoding compared to YHC (t(48) = -4.548, p < 0.001), (**Supplementary** Figure 1A). However, the two groups were not significantly different in Set Size at Maintenance (t(48) = - 0.936, p = 0.354). YHC had also a significantly lower Set Size Cost compared to EHC (t(47) = -3.391, p = 0.001). Bayesian independent sample test showed a BF_10_ = 523.1 for the Set Size at Encoding and BF_10_ = 23.28 for Set Size Cost, providing strong evidence for these effects.

Overall, performance at remembering three items simultaneously was much worse for older individuals compared to younger people, while when the WM load of encoding three items was distributed across the initial encoding and maintenance phases elderly participants were able to cope with the increased cognitive demand as well as younger ones.

#### 2.2.3 Impact of Filtering at Encoding and Maintenance

Both groups were able to efficiently filter out irrelevant items, shown by the Filtering rates being not significantly different from zero in both groups at Encoding and Maintenance (**Supplementary** Figure 1B). EHC and YHC were not significantly different in Filtering rate at Encoding. However, EHC had worse performances (higher values) compared to YHC in Filtering rate at Maintenance (U = 205, p = 0.038). Nevertheless, the Filtering rate Cost was not significantly different between the two groups. Moreover, the Bayesian independent sample test showed a BF_10_ = 1.047, providing no definitive evidence for a significant difference in Filtering rate at Maintenance between groups.

#### 2.2.4 Analysis of mixture model parameters

##### Precision

YHC had significantly higher Precision than EHC [F(1, 42) = 59.06, p < 0.001, η2 = 0.399]. There were no other significant main effects or interactions, suggesting the difference in precision reflects a pure ageing effect, regardless of task demands (Figure 2B**, panel a**).

##### Target detection

YHC showed significantly better target detection compared to EHC [F(1, 48) = 25.38, p < 0.001, η2 = 0.198], (Figure 2B**, panel b**). Across both groups, significantly more targets were detected in the filtering conditions than in SS3 conditions, indexed by a main effect of Instruction [F (1,48) = 10.01, p = 0.003, η2 = 0.031]. Furthermore, there was a main effect of Condition [F (1,48) = 6.64, p = 0.013, η2 = 0.011], with significantly fewer targets identified at Encoding than during Maintenance.

EHC performed worse than YHC when all three items needed to be remembered compared to when an item had to be filtered out, as shown by a significant two-way interaction between Instruction and Group [F (1,48) = 9.62, p = 0.003, η2 = 0.030]. Finally, there was a significant three-way Instruction x Condition x Group interaction [F (1,48) = 4.39, p = 0.041, η2 = 0.010], with EHC being worse at detecting targets when 3 items had to be remembered at Encoding compared to Maintenance. These results suggest that YHC and EHC found it more challenging to detect targets when all three items had to be encoded, and if the stimuli appeared at Encoding, with EHC paying the highest cost.

##### Misbinding

There was a significant main effect of Group [F(1, 43) = 15.29, p < 0.001, η2 = 0.103], and of Instruction [F(1, 43) = 6.74, p = 0.013, η2 = 0.038], with significantly more misbinding errors committed by both groups in the SS3 conditions compared to the filtering conditions (Figure 2B**, panel c**). Thus, even in healthy young individuals, misbinding rates were higher if three items had to be remembered compared to when one could be filtered out, suggesting that filtering can reduce WM resources consumption.

##### Guessing

A main effect of Group (F(1, 44) = 7.32, p = 0.010, η2 = 0.070], no main effects of either Instruction or Condition, but significant 2-way Instruction x Group (F(1, 44) = 4.329, p = 0.043, η2 = 0.016] and Instruction x Condition [F(1, 44) = 5.391, p = 0.025, η2 = 0.017] interactions were found (Figure 2B**, panel d**). This suggests that EHC guessed more than YHC, and more so in the more taxing SS3 conditions, independently of being presented during the Encoding or the Maintenance phase. Moreover, filtering during the Encoding phase led to more guessing in both groups, compared to when filtering had to be carried out during Maintenance.

Overall, Study 1 showed that both EHC and YHC were able to filter out distractors efficiently. EHC benefited from encoding three items non-simultaneously, and there was a weak effect of increased difficulty in Filtering during Maintenance compared to YHC. Overall, EHC made more errors compared to YHC, and this was due to worse performance across all types of errors: decline in memory Precision, reduced Target detection, increased Misbinding rates and increased Guessing. Filtering out an item was clearly beneficial in term of increased rates of target detection and reduced misbinding rates across the two groups. EHC also guessed more, especially when one item could not be filtered out. Memory precision was lower in EHC across all conditions and was the metric that showed the highest effect size, suggesting it might be a good marker of ageing, rather than reflecting task complexity or underlying filtering abilities.

## 3. Study 2 | Filtering in Alzheimer’s disease

### 3.1 Materials ad methods

#### 3.1.1 Participants

Twenty-eight patients with Alzheimer’s clinical syndrome, defined as per^61^, i.e., AD group, and 28 EHC (the same as Study 1) were recruited respectively from the Oxford Centre for Cognitive Disorders and through open day events. Participants underwent the Filtering at Encoding and Maintenance Task, a neuropsychological battery (**Table 1**), and a subset of patients (48/56) consented to perform a 3T MRI brain scan.

**Table 1.** | EHC and AD demographics and tests. Results are presented as mean, with standard deviation. According to normality of data, p-values were computed either by independent sample t-test or Mann-Whitney U test. Gender: M = male, F = Female. Education: years of full-time education. Handedness: Right-handed, Left-handed, Ambidextrous. n/a = not applicable. Addenbrooke’s Cognitive Examination-III (ACE-III), Digit Span (DS), Apathy Motivation Index (AMI), Beck’s Depression Inventory (BDI), Hospital Anxiety and Depression Scale (HADS), Snaith-Hamilton Pleasure Scale (SHAPS), Mood Scale (MS), Fatigue Severity Scale (FSS), Visual Analog Fatigue Scale (VAFS), Pittsburgh Sleep Quality Index (PSQI), World Health Organisation Five Well-Being Index (WHO-5), Cantril Ladder (CL) measure of life satisfaction. Disease duration: years since onset of symptoms.

|  | EHC | AD | p-value |
| --- | --- | --- | --- |
| Age | 70.36 (7.7) | 70.79 (9.5) | 0.854 |
| Gender (M/F) | 16/12 | 15/13 | 0.793 |
| **Education** | 17.75 (6.5) | 15.11 (5.5) | 0.036* |
| **Handedness (R/L/A)** | 22/6/0 | 23/3/2 | 0.859 |
| **ACE-III** | 97.82 (1.8) | 78.75 (11.8) | *< 0.001 |
| **DS** | 17.36 (3.6) | 16.04 (4.3) | 0.223 |
| **AMI** | 1.08 (0.4) | 1.7 (0.7) | *< 0.001 |
| **BDI** | 4.6 (4.5) | 10.86 (9.0) | *0.002 |
| **HADS** | 5.1 (3.0) | 8.69 (6.2) | *0.040 |
| **SHAPS** | 18.32 (4.7) | 20.79 (3.9) | *0.021 |
| **MS** | 1.29 (1.5) | 3.3 (3.5) | *0.005 |
| **FSS** | 2.8 (1.2) | 3.05 (1.5) | 0.591 |
| **VAFS** | 84.14 (21.1) | 67.86 (24.2) | *0.005 |
| **PSQI** | 4.36 (3.2) | 4.41 (3.2) | 0.905 |
| **WHO-5** | 78.0 (11.5) | 62.7 (20.8) | *0.009 |
| **CL** | 8.44 (1.2) | 7.79 (1.6) | 0.149 |
| **Disease duration** | - | 4.7 (3.1) | - |

The neuropsychological battery comprised measures of global cognition, verbal short-term memory, depression, apathy and fatigue. These included ACE-III, Digit Span (DS),^62^ Apathy Motivation Index (AMI),^63^ Beck’s Depression Inventory (BDI),^64^ Hospital Anxiety and Depression Scale (HADS),^65^ Snaith-Hamilton Pleasure Scale (SHAPS),^66^ Mood Scale (MS), i.e. the 15-Item Geriatric Depression Scale (GDS-15),^67^ Fatigue Severity Scale (FSS),^68^ Visual Analogue Fatigue Scale (VAFS),^69^ Pittsburgh Sleep Quality Index (PSQI),^70^ World Health Organisation Five Well-Being Index (WHO-5)^71^ and Cantril Ladder (CL).^72^ A summary of participants’ demographics and tests results is presented in **Table 1**. EHC and AD were not significantly different with respect to age, gender and handedness. All elderly healthy controls had global cognitive scores within the normal range, and none was significantly depressed.

#### 3.1.2 MRI acquisition and analysis

The MRI protocol and pipeline used to calculate head-size-corrected hippocampal volumes have been described elsewhere,^44^ and can be found in Supplementary materials.

#### 3.1.3 Statistical analysis

For MAE, Precision, Target detection, Misbinding, and Guessing, we performed 2 (**Instruction:** Filter and SS3) x 2 (**Condition:** Encoding and Maintenance) x 2 (**Group:** EHC, AD) mixed ANOVAs. We followed the same principles described in Study 1 in computing Set Size and Filtering rate metrics. Between-group differences were assessed through frequentist and Bayesian ANCOVAs using age, sex, education and handedness as covariates. The same principles were applied in Study 3.

### 3.2 Results

#### 3.2.1 Mean absolute error (MAE)

There was a main effect of Group [F (1,52) = 43.01, p < 0.001, η2 = 0.361], (Figure 3) and Instruction [F (1,52) = 16.54, p < 0.001, η2 = 0.022], with higher errors in the conditions where three items had to be remembered compared to when one item could be filtered out. There was no main effect of Condition, implying an overall equal number of errors at Encoding or Maintenance. Nevertheless, there was a significant Condition x Group interaction [(F (1,52) = 4.26, p = 0.044, η2 = 0.004], with AD performing worse at Maintenance, while EHC performing worse at Encoding. There was also a significant Instruction x Condition interaction [(F (1,52) = 11.79, p = 0.001, η2 = 0.010], with Filtering at Maintenance and Set Size at Encoding being challenging conditions compared to their counterparts Filtering at Encoding and Set Size at Maintenance.

**Figure 3.**
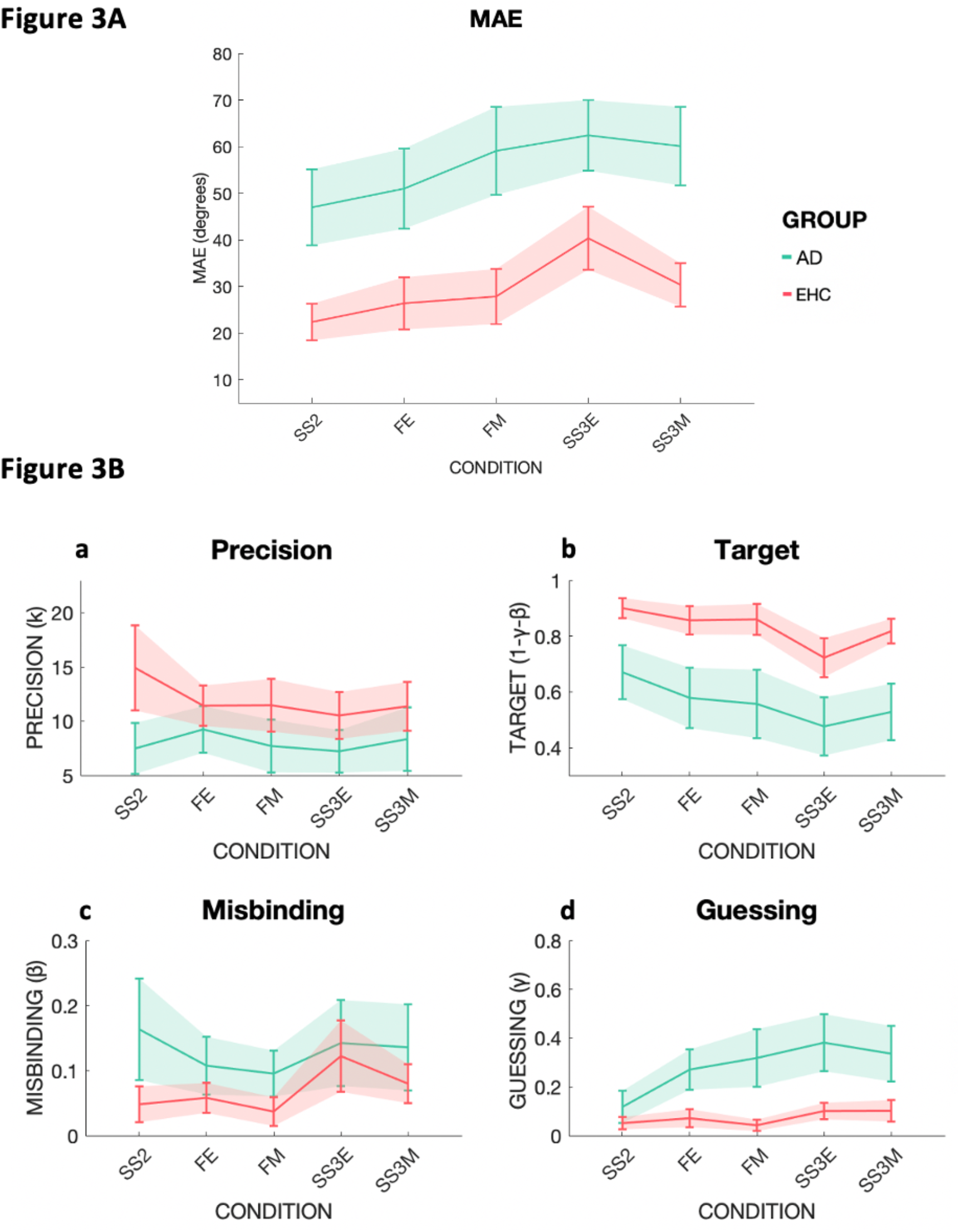
| Mean absolute error (MAE) and mixture model metrics in EHC and AD. Performance across the five different conditions: Set Size 2 (SS2), Filtering at Encoding (FE), Filtering at Maintenance (FM), Set Size three at Encoding (SS3E), Set Size three at Maintenance (SS3M). The two groups are shown as EHC in coral red, AD in green. **3A:** On the Y-axis MAE in degrees. Note in particular the differential effect of Filtering at Maintenance (FM) compared to Filtering at Encoding (FE) within the AD group compared to within the EHC group. Further, in AD, FM performance (when a to-be-ignored distractor was presented during maintenance) was equivalent to that Set Size three at Maintenance (SS3M, when the new item had to be retained). **3B: a |** Precision as concentration parameter κ. **b |** Target probability. **c |** Misbinding probability. **d |** Guessing probability.

Overall, AD patients performed worse than EHC, and regardless of the diagnosis, participants made more errors when three items had to be remembered compared to when one had to be filtered out. Across both groups, filtering seemed to be more taxing during Maintenance, while remembering three items was more difficult if they appeared simultaneously at Encoding. AD patients experienced greater difficulty at Maintenance, whereas EHC performed worse at Encoding.

#### 3.2.2 Impact of Set Size at Encoding and Maintenance

AD patients showed a greater Set Size effect at Maintenance [F (1,49) = 4.35, p = 0.042] compared to EHC, but no difference in Set Size at Encoding, or their Cost (**Supplementary** Figure 2A). The Bayesian ANCOVA showed low evidence for this effect (BF_M_ = 3.160). This low effect is also in line with the absence of a difference in Set Size Cost.

#### 3.2.3 Impact of Filtering at Encoding and Maintenance

There was no difference in Filtering rate at Encoding between the two groups, but groups were different in Filtering rate at Maintenance [F (1,48) = 8.94, p = 0.004] and Filtering rate Cost [F (1,48) = 4.63, p = 0.036], with AD having higher Filtering rates at Maintenance and lower Filtering rate Cost, indicative of worse performances at Maintenance (**Supplementary** Figure 2B). The Bayesian ANCOVA showed a BF_M_ = 5.762 for Filtering rate at Maintenance and BF_M_ = 3.324 for Filtering rate Cost, therefore providing evidence for a significant difference.

#### 3.2.4 *Analysis of mixture model parameters* Precision

AD patients had lower Precision compared to EHC [F (1,39) = 4.34, p = 0.044, η2 = 0.039] (Figure 3B**, panel a**). There were no main effects of Instruction or Condition, nor any significant interactions. Therefore, AD had lower precision compared to EHC regardless of the condition.

##### Target detection

For Target detection, a main effect of Group [F (1,52) = 36.66, p < 0.001, η2 = 0.291], with AD patients exhibiting worse performance compared to EHC, was found (Figure 3B**, panel b**). There was also a main effect of Instruction [F (1,52) = 10.20, p = 0.002, η2 = 0.021], with less targets detected in SS3 than in the filtering conditions. While AD patients were worse at detecting targets compared to EHC, both groups benefitted from filtering one item out.

##### Misbinding

There was a main effect of Group [F (1,52) = 7.87, p = 0.007, η2 = 0.047], with higher misbinding rates in AD patients compared to EHC (Figure 3B**, panel c**) and a main effect of Instruction [F (1,52) = 6.02, p = 0.018, η2 = 0.029], with more misbinding in SS3 than in the filtering conditions for both groups.

##### Guessing

A main effect of Group [F (1,48) = 31.27, p < 0.001, η2 = 0.241], with AD patients performing worse than EHC (Figure 3B**, panel d**), was found.

In the AD group there was no correlation between disease duration and filtering abilities, working memory capacity or any other WM metric.

Overall, in line with Study 1, Study 2 showed that filtering out an item was clearly beneficial in term of increased rates of Target detection and reduced Misbinding across the two groups but was unable to increase memory Precision. Filtering at Maintenance was worse in AD patients compared to EHC, with AD patients showing similar performances in FM as in the SS3M condition (Figure 3). AD patients showed impairment across all multiple metrics compared to EHC: reduced memory Precision and Target detection, increased Misbinding and Guessing. However, in this case Target detection and Guessing, and not memory Precision as in healthy ageing, showed the strongest effect size in group comparisons.

## 4. Study 3 | Filtering in Parkinson’s disease

### 4.1 Materials and methods

#### 4.1.1 Participants

Twenty-eight patients with idiopathic Parkinson’s Disease, diagnosed as per^73^, were recruited from the Cognitive Disorder Clinic and the Oxford Parkinson’s Disease Centre Discovery Cohort, based at the John Radcliffe Hospital in Oxford. Their performance was compared to the EHC group of Study 1 and 2. We recruited only PD patients with no evidence of cognitive impairment (ACE-III total score > 88/100), to avoid potential confounding factors in results’ interpretation. Nevertheless, further studies including a wider range of cognitively intact PD patients, PD with mild cognitive impairment and PD patients with dementia would be advisable. PD patients were tested in ‘ON’ state, i.e. within 3 hours of their usual dopaminergic medications. Participants underwent the same assessments described in Study 2, and a subset (43/56) consented to MRI brain scanning. In addition, we collected data on Unified Parkinson’s Disease Rating Scale (UPDRS) scores and total Levodopa dose, to investigate whether disease stage or dopaminergic medications could impact performance. Participants’ demographics and test scores are presented in **Table 2**. PD patients had lower ACE scores, but still within normal ranges (above 88/100 total).

**Table 2.**
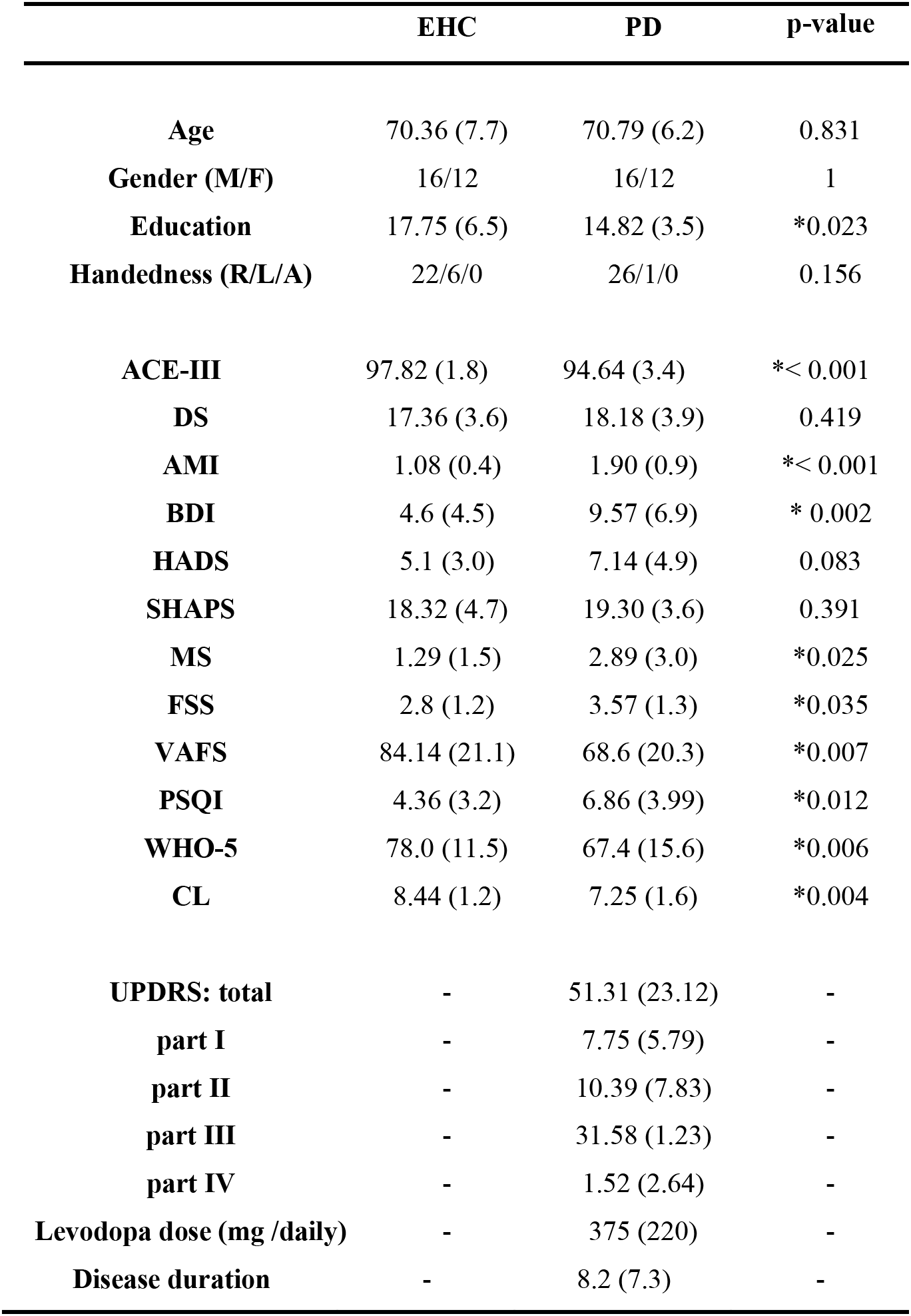
| EHC and PD demographics and tests. Results are presented as mean and SD in brackets. According to normality of the data, p-values were computed either by independent sample t-test or Mann-Whitney U test. Gender: M = male, F = Female, shown as male/female ratio. Education is expressed in years of full-time education. Handedness: Right-handed, Left-handed, Ambidextrous. n/a = not applicable. Addenbrooke’s Cognitive Examination-III (ACE-III), Digit Span (DS), Apathy Motivation Index (AMI), Beck’s Depression Inventory (BDI), Hospital Anxiety and Depression Scale (HADS), Snaith-Hamilton Pleasure Scale (SHAPS), Mood Scale (MS), Fatigue Severity Scale (FSS), Visual Analog Fatigue Scale (VAFS), Pittsburgh Sleep Quality Index (PSQI), World Health Organisation Five Well-Being Index (WHO-5), Cantril Ladder (CL) measure of life satisfaction, Unified Parkinson’s Disease Rating Scale (UPDRS). Disease duration: years since onset of symptoms.

|  | **EHC** | **PD** | **p-value** |
| --- | --- | --- | --- |
| **Age** | 70.36 (7.7) | 70.79 (6.2) | 0.831 |
| **Gender (M/F)** | 16/12 | 16/12 | 1 |
| **Education** | 17.75 (6.5) | 14.82 (3.5) | *0.023 |
| **Handedness (R/L/A)** | 22/6/0 | 26/1/0 | 0.156 |
| **ACE-III** | 97.82 (1.8) | 94.64 (3.4) | *< 0.001 |
| **DS** | 17.36 (3.6) | 18.18 (3.9) | 0.419 |
| **AMI** | 1.08 (0.4) | 1.90 (0.9) | *< 0.001 |
| **BDI** | 4.6 (4.5) | 9.57 (6.9) | * 0.002 |
| **HADS** | 5.1 (3.0) | 7.14 (4.9) | 0.083 |
| **SHAPS** | 18.32 (4.7) | 19.30 (3.6) | 0.391 |
| **MS** | 1.29 (1.5) | 2.89 (3.0) | *0.025 |
| **FSS** | 2.8 (1.2) | 3.57 (1.3) | *0.035 |
| **VAFS** | 84.14 (21.1) | 68.6 (20.3) | *0.007 |
| **PSQI** | 4.36 (3.2) | 6.86 (3.99) | *0.012 |
| **WHO-5** | 78.0 (11.5) | 67.4 (15.6) | *0.006 |
| **CL** | 8.44 (1.2) | 7.25 (1.6) | *0.004 |
| **UPDRS: total** | - | 51.31 (23.12) | - |
| **part I** | - | 7.75 (5.79) | - |
| **part II** | - | 10.39 (7.83) | - |
| **part III** | - | 31.58 (1.23) | - |
| **part IV** | - | 1.52 (2.64) | - |
| **Levodopa dose (mg /daily)** | - | 375 (220) | - |
| **Disease duration** | - | 8.2 (7.3) | - |

### 4.2 Results

#### 4.2.1 Mean absolute error (MAE)

There was no significant group difference between EHC and PD patients in MAE (Figure 4A). However, there was a main effect of Instruction [F (1,49) = 27.07, p < 0.001, η2 = 0.079], with more targets detected filtering conditions. There was an Instruction x Condition interaction [F (1,49) = 8.12, p = 0.006, η2 = 0.016], suggesting remembering three items was more difficult at Encoding.

**Figure 4.**
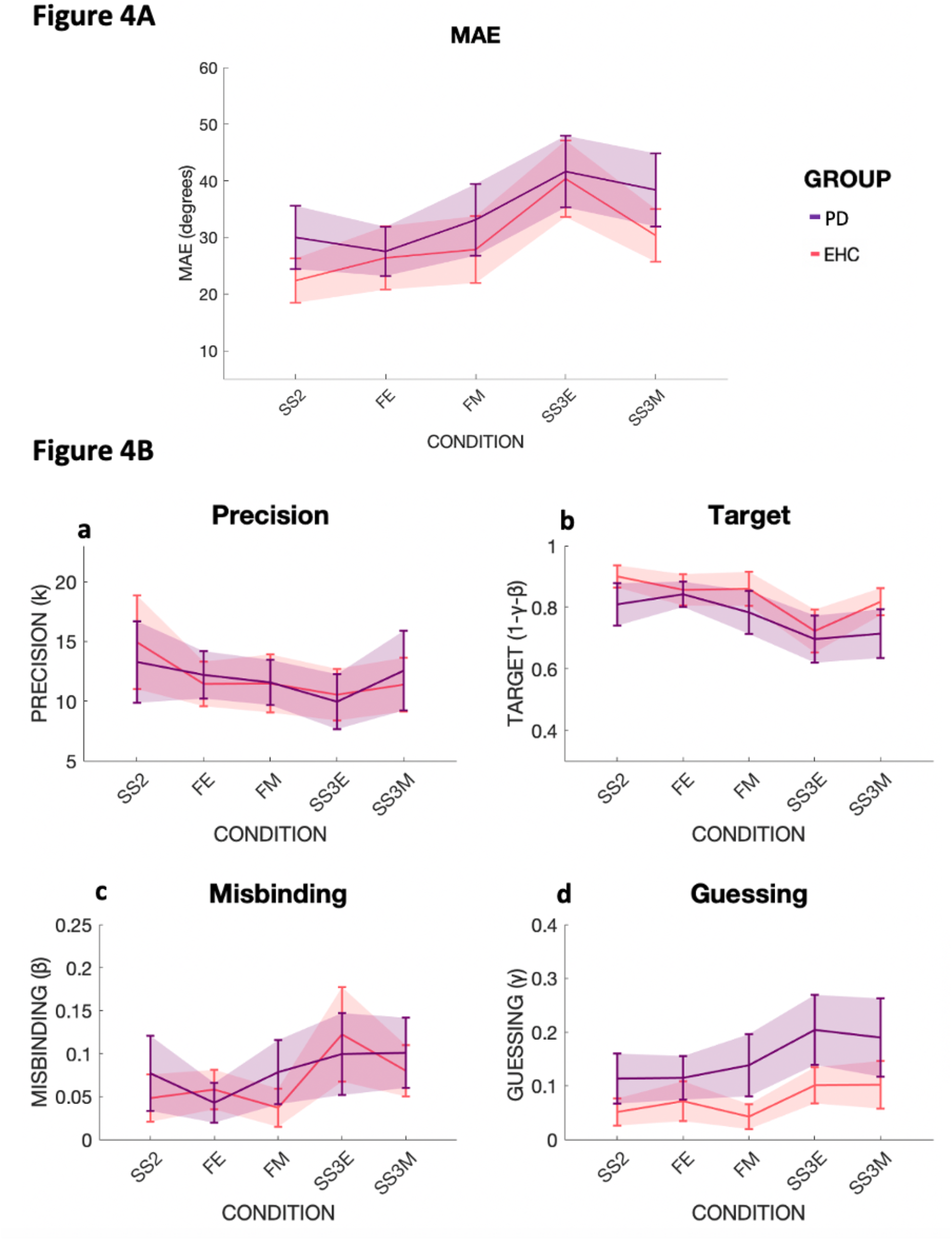
| Mean absolute error (MAE) and mixture model metrics in EHC and PD. Performance across the five different conditions: Set Size 2 (SS2), Filtering at Encoding (FE), Filtering at Maintenance (FM), Set Size three at Encoding (SS3E), Set Size three at Maintenance (SS3M). Different participant groups are labeled with different colors, i.e. EHC in coral red, PD in violet. **4A:** On the Y-axis, MAE in degrees for the two groups. **4B: a |** Precision as concentration parameter κ. **b |** Target probability. **c |** Misbinding probability. **d |** Guessing probability.

#### 4.2.2 Set Size and Filtering strategies

There was no significant difference in Set Size at Encoding, Maintenance, or their Cost, nor in Filtering rate at Encoding, Maintenance and their Cost, among the two groups.

#### 4.2.3 *Analysis of mixture model parameters* Precision

No main effect of Group, Instruction or Condition, but a significant Instruction x Condition interaction [F (1,38) = 4.58, p = 0.039, η2 = 0.022] was found, suggesting that overall participants were more precise during Set Size three at Maintenance rather than Set Size three at Encoding (Figure 4B**, panel a**). Therefore, memory Precision does not seem to be affected in PD.

##### Target detection

There were no main effects of Group, or Condition, but a main effect of Instruction [F (1,49) = 29.60, p < 0.001, η2 = 0.090], with less target detected in conditions where three items needed to be remembered compared to when one item could be filtered out (Figure 4B**, panel b**).

##### Misbinding

No main effects of Group, or Condition, but a main effect of Instruction [F (1,46) = 7.72, p = 0.008, η2 = 0.046] was seen, with misbinding occurring more frequently when three items need to be remembered (Figure 4B**, panel c**).

##### Guessing

There was a main effect of Group [F (1,45) = 9.43, p = 0.004, η2 = 0.074], with PD guessing significantly more than EHC, and a main effect of Instruction [F (1,45) = 8.50, p = 0.006, η2 = 0.037], with more Guessing occurring in the conditions when all three items had to be remembered (Figure 4B**, panel d**).

In the PD group, we found no correlation between UPDRS, Levodopa dose or disease duration and filtering abilities, working memory capacity or any of the WM metrics.

In agreement with Study 1 and 2, Study 3 showed that filtering out an unwanted item improved Target detection, and reduced Misbinding, while having no effect on memory Precision. The key result was that PD patients exhibited a selective increase in Guessing compared to EHC, but otherwise had comparable performances in terms of WM filtering efficiency and capacity, memory Precision, Target detection and Misbinding rates. Another novel finding compared to Study 1 and 2, is that in Study 3 filtering out an item was beneficial also to reduce Guessing rates in EHC and PD if taken together.

## 5. Transdiagnostic analysis of cognitive measures and hippocampal volumes

We next investigated the interplay between WM capacity, filtering abilities, different sources of error derived from the Mixture Model, and hippocampal volumes using a transdiagnostic approach. Cumulative metrics for each Mixture Model variable were calculated by averaging the performance across all five conditions (SS2, FE, FM, SS3E, SS3M). Correlations between variables were computed using a generalized linear model, with age, sex, handedness and education included as covariates of no interest.

### 5.1 Relationship between hippocampal volumes and filtering ability

Across groups, *Filtering at Maintenance* was significantly negatively correlated with whole hippocampal volume (WHV) (r = –0.404 p = 0.002), (Figure 5a, Figure 6). Figure 6 displays the relationship between Filtering at Maintenance and hippocampal volumes, with participants with higher Filtering at Maintenance (on the right side), having lower hippocampal volumes (bottom of the graph). There was no significant correlation between hippocampal volume measures with Filtering at Encoding or with Set Size, either at Encoding or Maintenance. Thus, Filtering at Maintenance was strongly related to hippocampal volume, which is often used as an important marker of AD (as age effects were regressed out in this analysis). To validate this result, we looked at whether Filtering at Maintenance was correlated with whole brain volume, and there was no such correlation (r = –0.116, p = 0.392).

**Figure 5.**
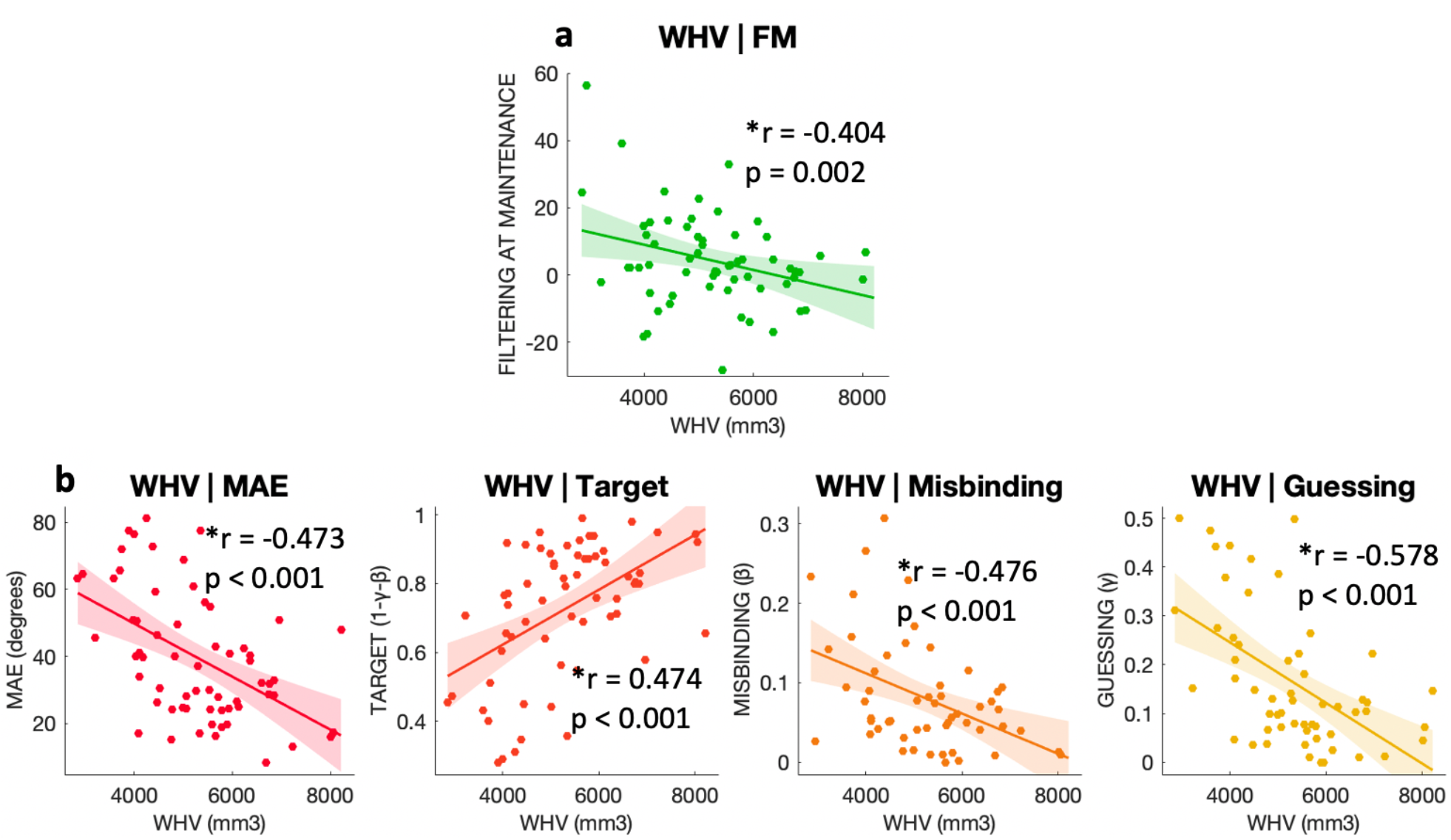
| Mixture model parameters correlations with hippocampal volumes. **a** | Correlation between Filtering rate at Maintenance and whole hippocampal volume (WHV). **b |** Correlation between Mixture Model metrics and whole hippocampal volume. On the X-axis the volume in mm^3^ of the whole hippocampus. In **b**, the Y-axis represent from left to right respectively MAE in degrees, probabilities of Target detection, Misbinding and random Guessing.

**Figure 6.**
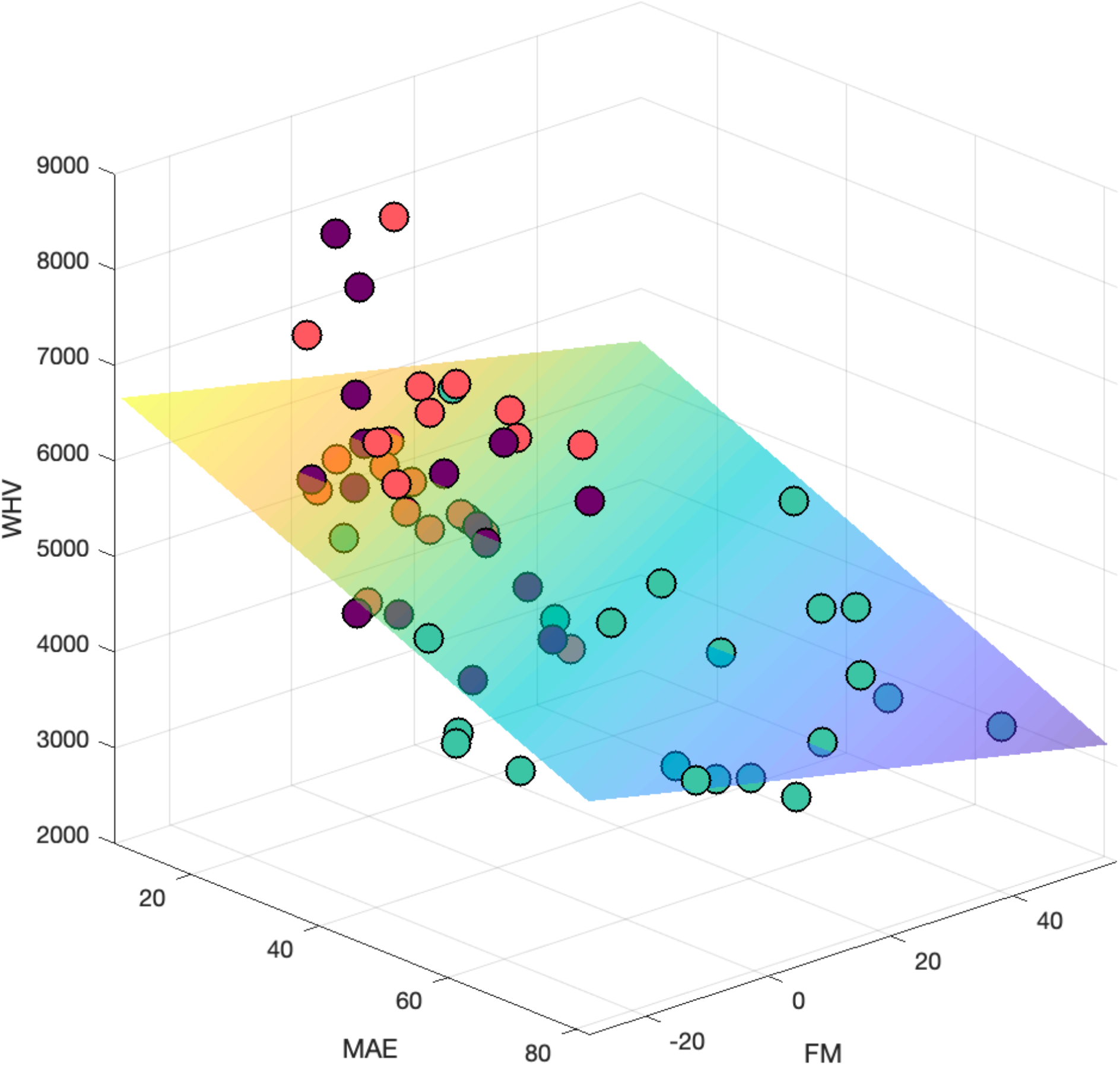
| Filtering and MAE stratified by WHV and diagnosis. FM on the X-axis, MAE on the Y-axis, hippocampal volumes (WHV) on the Z-axis. Subjects with higher WHV (top of the image) have lower MAE (back of the image), lower Filtering at Maintenance scores (on the left) and are predominantly represented by elderly healthy subjects (in coral red), and PD patients (in violet). On the other end of the spectrum, AD patients (in green) have higher Filtering at Maintenance scores (on the right), higher MAE (front of the image), and have lower hippocampal volumes (bottom of the image).

### 5.2 Relationship between hippocampal volume and other WM performance metrics

WHV was negatively correlated with MAE (WHV: r = –0.473, p < 0.001), Misbinding (WHV: r = –0.476, p < 0.001) and Guessing (WHV: r = –0.578, p < 0.001) and positively correlated with Target detection (WHV: r = 0.474, p < 0.001), (Figure 5b, Figure 6 for MAE). Figure 6 shows how higher MAE is negatively correlated with hippocampal volumes, as the regression plane goes from higher hippocampal values (top if the image) to lower values (bottom) with increasing MAE values (from back to front of the image on the Y-axis). No association between hippocampal volume and Precision was found (WHV: r = -0.057, p = 0.710). In this case, both MAE (r = –0.319, p = 0.014), Target detection (r = 0.310, p = 0.017) and Guessing (r = -0.335, p = 0.015) were correlated with whole brain volume, while Misbinding was not.

We then performed a stepwise regression analysis to determine which metric was the best predictor of hippocampal volume, with WHV as dependent variable, MAE, Misbinding, Guessing, and Filtering at Maintenance as covariates. Target detection was not included for avoiding collinearity in multiple regression, which arises from how Target, Misbinding and Guessing probabilities are calculated. Overall, WHV was better explained by Guessing (r = 0.526, p < 0.001).

## 6. Discussion

This study, using a novel delayed reproduction task, compared WM filtering performance at encoding and maintenance in healthy ageing, AD and PD.

Previous evidence suggests that filtering abilities are tightly related to higher WM capacity.^20,74^ Our data support this view, as shown by Figure 6, where there is a positive correlation between mean absolute error and filtering at maintenance. Elderly controls showed preserved filtering abilities if the distractor was presented at Encoding (Figure 2A). However, their ability to filter out a distractor during the Maintenance period might not be as efficient, as suggested by the small age-related effect in decline in Filtering at Maintenance (**Supplementary** Figure 1B). This is in line with previous evidence of an age-dependent decline in performance during maintenance,^20^ but in our case, the Bayesian analysis did not provide strong evidence to draw a firm conclusion. This might be due to our elderly participants being thoroughly screened for depression, apathy and cognitive impairment, which might lead to a potential inflation of their cognitive abilities compared to large, community-based studies.

AD patients on the other hand showed much clearer WM deficits if they had to filter out an irrelevant distractor during the Maintenance phase (Figure 3A**, Supplementary** Figure 3B). AD patients have been found to struggle to prioritize relevant over irrelevant visual information across different tasks where an optimal inhibitory control is required to avoid unwanted attentional capture^75,76,77,78^. Our study was able to show not only that filtering abilities are impaired in AD patients, but that their deficit is specifically limited to the maintenance phase.

In contrast, PD patients showed excellent filtering ability both at Encoding and Maintenance. All our PD patients were tested in an ON Levodopa state (when dopaminergic medications are effective). Other studies have reported an impaired filtering ability in PD patients OFF Levodopa,^55^ so it is possible that there is a beneficial effect of dopamine replenishment on ignoring or filtering in PD.^17^ The integrity of WM maintenance processes in PD concurs with previous data on visual pattern tasks that likely require both object and spatial processing,^79^ and our results suggest that PD patients are able to manipulate information successfully during the maintenance period, unlike AD patients. Since we did not specifically test for a delay effect, we cannot exclude that at longer delay PD patients could also suffer from delay-dependent declines in maintenance performances as previous evidence suggests.^80^

Overall, healthy elderly participants performed worse compared to younger participants and were specifically impaired when three items had to be encoded simultaneously (Figure 2, **Supplementary** Figure 1A), in line with a well-known age-related decline in WM capacity.^18,29^ AD patients showed overall lower performances compared to elderly controls. Unlike the latter, however, they seemed to cope well with increased Set Size at Encoding (Figure 3**, Supplementary** Figure 2A), but were mildy impaired during the Maintenance phase, as reflected by the weak Set Size effect at Maintenance.

Mixture Model analysis revealed that healthy ageing is associated with different types of errors: reduced memory Precision, decreased Target detection, increased Misbinding and increased Guessing (Figure 2B). But while filtering out an item was beneficial in increasing the numbers of detected targets, and reducing Misbinding, it was not able to help maintaining a high memory Precision, which declined invariably across all conditions in elderly subjects (Figure 2B**, panel a**). Filtering out a distractor was also beneficial in reducing Guessing, and that was true specifically for EHC, with YHC not showing such effect. A possible interpretation is that the task was more taxing for the EHC, who had to deploy a cognitive strategy to free up space in WM. These results support the idea that memory Precision could be considered as a marker of ageing, as previously reported by other groups.^15,18,19,20^ In this study, we were also able to show that not only memory Precision declines with ageing, but also that AD patients have a further drop in memory precision compared to age-matched healthy controls, albeit of a much smaller magnitude, while this does not occur in PD patients.

Another memory feature that seems to decline with ageing is the ability to correctly identify a target.^15,81,82^ Our results are consistent with this view since a clear decline in Target detection was observed in elderly compared to younger controls across all conditions (Figure 2B**, panel b**). There was also a synergic detrimental effect in ageing, as reflected by a significant three-way Instruction x Condition x Group interaction in Study 1, suggesting that elderly controls were worse at detecting targets when three items had to be remembered, and when this had to be accomplished during the Encoding phase. AD patients were even less likely to correctly identify a target compared to age-matched controls (Figure 3B**, panel b**). This supports previous findings of reduced correct object identity recognition in familial AD^35^ as well as sporadic, late onset AD.^44, 83^

In line with previous data,^15,18^ Misbinding increased in healthy ageing, particularly if more items had to be remembered (Figure 2B**, panel c**). Misbinding was further increased in AD patients compared to healthy controls (Figure 3B), unlike in PD patients (Figure 4B). Previous data show that both familial^35^ and sporadic late onset AD patients^12,44^ exhibit increased rates of Misbinding. Whilst confirming that object binding is a feature that is frail in healthy ageing, and more so, in AD, we also found that filtering out an item decreases Misbinding rates in EHC, AD and PD.

Lastly, Guessing rates were higher in healthy ageing, AD and PD patients (Figure 2B**, 3B, 4B, panel d**), and it was the only metric which could differentiate between PD patients and EHC. This supports previous evidence showing that Guessing is primarily affected in PD, but can be increased in AD patients.^44^ Filtering out an item was able to help Guessing less in EHC but not YHC, in Study 1, was ineffective in AD patients in Study 2, and was beneficial in EHC and PD in Study 3. Previous studies have specifically linked increased misbinding to hippocampal dysfunction.^45,47,84,85,86^ Our results support this view (Figure 5b), and the lack of correlation with whole brain volume, unlike other metrics such as mean absolute error or target detection, also reinforces the idea that this metric reflects hippocampal integrity.

In our dataset Guessing was the best predictor across the WM examined. Notably, our sample consisted of an heterogeneous population, comprising also PD patients, which are more prone to guessing, as well more cognitively impaired AD patients compared to similar studies.^35,46^ For misbinding errors to occur, at least a partial memory of the object (its identity) must be retained, while the memory of its location might be lost. When no mnemonic trace is left, Guessing remains the only viable option, and therefore its rates might be higher in more cognitively impaired populations.

While higher Target detection was associated with higher hippocampal volumes, this was not the case for memory Precision. Independent findings suggest that the parietal cortex rather than the hippocampus could be primarily involved in maintaining information with high detail of precision over time,^37,87^, and therefore we cannot exclude that this could be the reason underlying the lack of structural associations with Precision in our sample.

Our data demonstrate also that filtering ability during maintenance – but not encoding – is related to hippocampal volume (Figure 5a**, 6**), suggesting that the hippocampus might protect the contents of WM, once encoded. This is further supported by the lack of correlation between Filtering at Maintenance and whole brain volume, pointing towards a specific contribution of the hippocampus in WM filtering during maintenance.

## 7. Conclusion

Elderly participants and PD had relatively preserved filtering abilities both at encoding and during maintenance. In AD, however, there were significant filtering deficits specifically when the distractor appeared during maintenance. Healthy ageing was associated with a more prominent decline in memory precision. In AD patients higher guessing and lower target detection were the leading sources of memory error while PD patients showed only higher guessing rates. Finally, hippocampal volume correlated significantly with filtering ability during maintenance – but not at encoding, as well as other mixture model metrics of WM performance, providing further evidence for the role of the hippocampus in WM.

## Data Availability

The fully anonymized dataset of this study can be found at: https://osf.io/3s6ze/.

## Acknowledgements and Funding

This work was supported by a Wellcome Trust Principal Research Fellowship to Masud Husain and by the NIHR Oxford Biomedical Research Centre. This research was funded in whole, or in part, by the Wellcome Trust [Grant Number: 206330/Z/17/Z]. For the purpose of Open Access, the author has applied a CC BY public copyright licence to any Author Accepted Manuscript version arising from this submission.

## Author contributions

**Sofia Toniolo**: Conceptualization, Methodology, Formal analysis, Investigation, Resources, Data curation, Writing - Original Draft, Writing - Review & Editing, Visualization, Project administration. **Robert Udale**: Conceptualization, Methodology, Software, Validation, Formal analysis, Resources. **Verena Svenja Klar**: Data curation. **Maria Raquel Maio**: Data curation. **Bahaaeddin Attaallah**: Data curation. **George Tofaris**: Resources. **Michele Tao-Ming Hu**: Resources. **Sanjay George Manohar**: Resources, Supervision. **Masud Husain**: Resources, Writing - Review & Editing, Supervision, Funding acquisition.

