## Supplementary materials for "WORKING MEMORY FILTERING AT ENCODING AND MAINTENANCE IN HEALTHY AGEING, ALZHEIMER’S AND PARKINSON’S DISEASE"

#### Study 1

##### Demographics

| | YHC | EHC | p-value |
|---|---|---|---|
| **Age** | 24.8 (5.1) | 70.36 (7.7) | *< 0.001 |
| **Gender (M/F)** | 10/18 | 16/12 | 0.113 |
| **Education** | 17.36 (4.2) | 17.75 (6.5) | 0.610 |
| **Handedness (R/L)** | 26/2 | 22/6 | 0.134 |
| **ACE-III** | - | 97.82 (1.8) | - |

**Supplementary Table 1 | Demographics for young and elderly healthy controls**

Values for age, education handedness and ACE-III are presented as mean (standard deviation). Gender: M = male, F = Female, is presented as male/female ratio. Education is shown as years of full-time education. Handedness: Right-handed, Left-handed. ACE-III: Addenbrooke's Cognitive Examination-III scores. Between-group differences are shown as p-values. According to the normality of the data, between-group differences in demographic data were assessed either with t-test, Mann-Whitney U test or $\chi^2$ test.

##### Filtering at Encoding and Maintenance Task parameters

Stimuli were displayed on a 24-inch LCD computer at a viewing distance of 60cm. The orientations of the arrows were located along the boundary of an imaginary circle, with radius 8° in visual angle, centered on the cardinal center of the screen. Each arrow was 1.4° x 0.6° visual angle in size and pointing in random directions over 360 degrees. Their orientations were random but constrained to be separated by at least 20 degrees of one another. Stimulus colors were red (RGB: [255, 0, 0]), green (RGB: [0, 255, 0]) or blue (RGB: [0, 0, 255]). Which arrow had to be ignored on a given trial was chosen at random. All stimuli were presented on a grey background (RGB: [171, 171, 171]). The arrow used to probe a response was black (RGB: [0, 0, 0]). Participants performed a total of 120 trials (5 conditions x 24 trials per condition), divided into 12 blocks of 10 trials each. Trials from all 5 conditions were interleaved within each block.

**Model 1**

The probability of responding with the reported orientation ($\hat{\theta}$) is described by the following equation:

$$p(\hat{\theta}) = (1 - \gamma - \beta)\Phi_{\kappa}(\hat{\theta} - \theta) \; + \gamma\frac{1}{2\pi} + \beta\frac{1}{m}\sum_{i}^{m}\Phi_{\kappa}(\hat{\theta} - \theta_i)$$

where θ is the true orientation of the target item, $\hat{\theta}$ the orientation reported by the participant, and $\varphi_k$ is the von Mises distribution (the circular analogue of the Gaussian) with mean zero and concentration parameter κ. The probability of reporting the correct target item is given by $1 - \gamma - \beta$. The probability of mistakenly reporting a non-target item is given by $\beta$, where {$\theta_1$, $\theta_2$, $\theta_m$} are the orientations of the $m$ non-target items. The probability of responding randomly is given by $\gamma$.

**Supplementary Figure 1A**

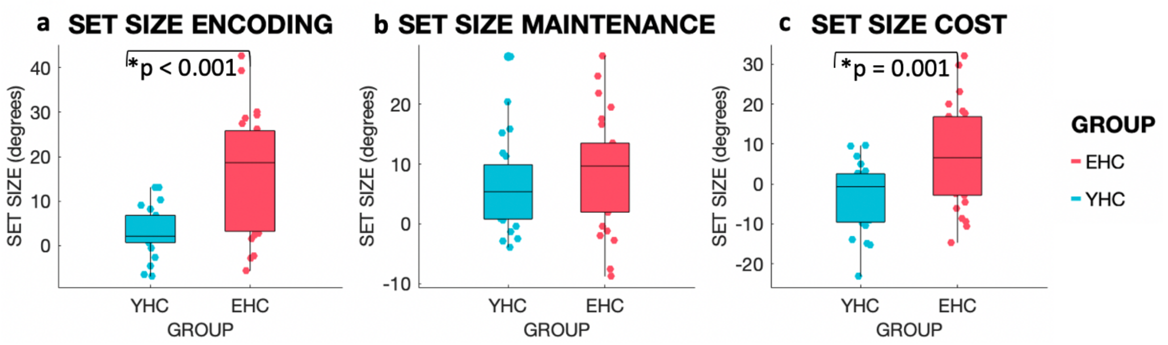


**Supplementary Figure 1B**

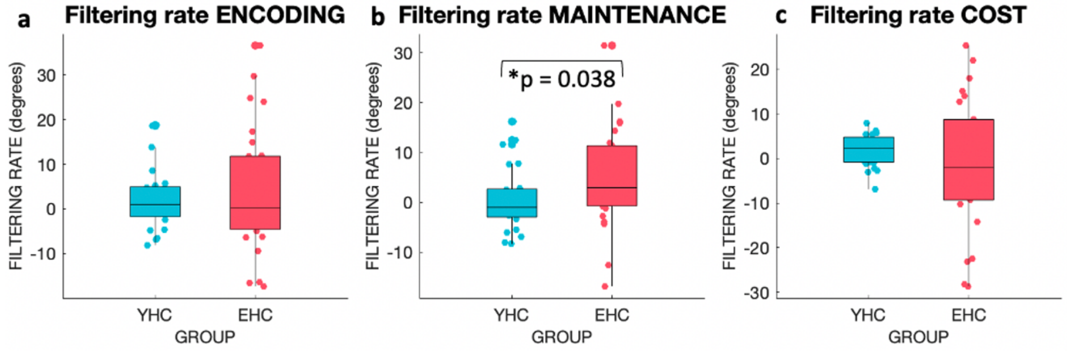


**Supplementary Figure 1 | Set size and filtering rate at encoding and maintenance in YHC and EHC**

**1A**: Set Size at Encoding, Maintenance and their Cost in YHC and EHC. Panel a: Set Size at Encoding (MAE SS3E - MAE SS2), panel b: Set Size at Maintenance (MAE SS3M- MAE SS2), panel c: Set Size Cost (Set Size at Encoding - Set Size at Maintenance). **1B:** Filtering rate at Encoding, Maintenance and their Cost in YHC and EHC. Filtering rate is given in degrees. Panel a: Filtering rate at Encoding (FE – SS2), panel b: Filtering rate at Maintenance (FM – SS2), panel c: Filtering Rate Cost (Filtering rate at Encoding - Filtering rate at Maintenance). Only statistically significantly p-values are shown.

### Study 2

#### MRI acquisition and analysis

T1-weighted volumetric MR brain images were acquired on a 3T Siemens Magnetom Verio syngo scanner using a magnetisation prepared rapid gradient echo (MPRAGE) protocol acquired in sagittal orientation (TR = 2000 msec, TE = 1.94 msec, TI = 880 msec, Flip angle = 8 degrees, FOV read = 256 mm, Voxel size = 1.0 x 1.0 x 1.0 mm). All images were reviewed by a trained neurologist to exclude the presence of remarkable macroscopic brain abnormalities not compatible with the original diagnosis. Hippocampal volumes (HV) were estimated using FSL-FIRST. For each participant, left, and right HV were calculated, and bilateral HV, or whole hippocampal volume (WHV) was computed. We also calculated whole brain volumes for each subject. Moreover, we derived the scaling factor to be used as normalisation for head size using SIENAX, after brain extraction and affine-registered to MNI152 space. We subsequently computed the head size corrected values for whole brain volumes and WHV, using the scaling factor derived from SIENAX. When referring to WHV and whole brain volumes throughout the article, only head-size corrected volumes have been used. Images were carefully visually inspected after each processing step.

#### Supplementary Figure 2A

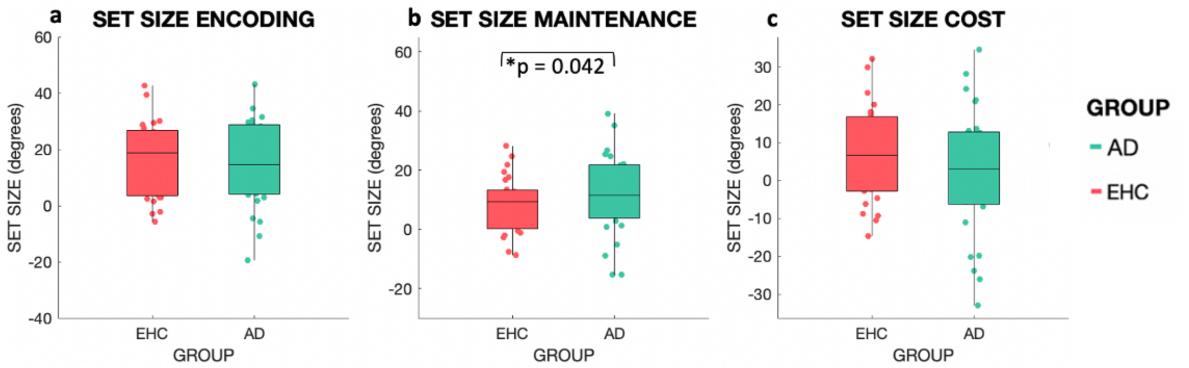


#### Supplementary Figure 2B

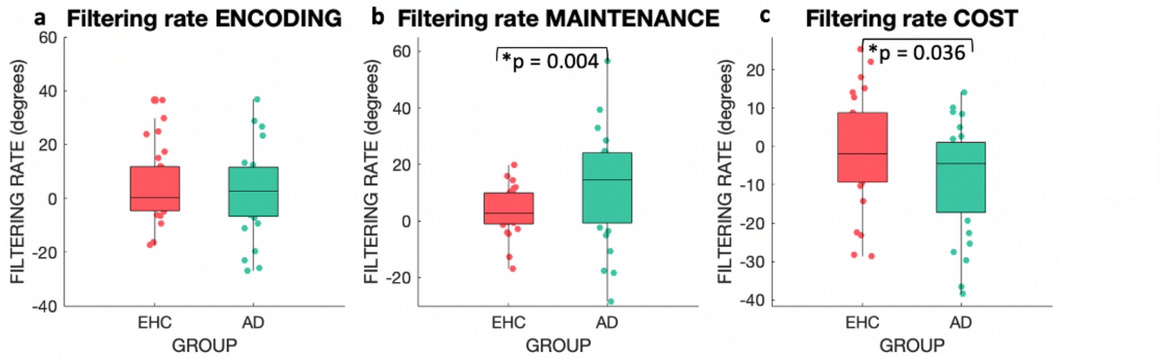


**Supplementary Figure 2 | Set size and filtering rate at encoding and maintenance in EHC and AD**

On the X-axis different participant groups are shown, labeled with different colors, i.e. EHC in coral red, AD in green. **2A: a** | Set Size at Encoding (MAE SS3E - MAE SS2). **b** | Set Size at Maintenance (MAE SS3M- MAE SS2). **c** | Set Size Cost (Set Size at Encoding - Set Size at Maintenance). **2B**: **a** | Filtering rate at Encoding (FE – SS2). **b** | Filtering rate at Maintenance (FM – SS2). **c** | Filtering Rate Cost (Filtering rate at Encoding - Filtering rate at Maintenance). Only statistical significantly differences are shown.
